# Increasing representation and diversity in health research: A protocol of the MYHealth Research Training Program for high school students

**DOI:** 10.1101/2023.02.02.23285366

**Authors:** Samantha A. Chuisano, Jane Rafferty, Alison Allen, Tammy Chang, Matthew Diemer, Kara Harris, Lisa M. Vaughn, Daphne C. Watkins, Melissa DeJonckheere

## Abstract

**Background:** Despite decades of calls for increased diversity in the health research workforce, disparities exist for many populations, including Black, Indigenous, and People of Color individuals, those from low-income families, and first-generation college students. To increase representation of historically marginalized populations, there is a critical need to develop programs that strengthen their path toward health research careers. High school is a critically important time to catalyze interest and rebuild engagement among youth who may have previously felt excluded from science, technology, engineering, and mathematics (STEM) and health research careers.

**Methods:** The overall objective of the MYHealth program is to engage high school students in a community-based participatory research program focused on adolescent health. Investigators will work alongside community partners to recruit 9^th^ through 12^th^ graders who self-identify as a member of a group underrepresented in STEM or health research careers (e.g., based on race and ethnicity, socioeconomic status, first generation college student, disability, etc.). MYHealth students are trained to be co-researchers who work alongside academic researchers, which will help them to envision themselves as scientists capable of positively impacting their communities through research. Implemented in three phases, the MYHealth program aims to foster a continuing interest in health research careers by developing: 1) researcher identities, 2) scientific literacy, 3) scientific self-efficacy, and 4) teamwork and leadership self-efficacy. In each phase, students will build knowledge and skills in research, ethics, data collection, data analysis, and dissemination. Students will directly collaborate with and be mentored by a team that includes investigators, community advisors, scientific advisors, and youth peers.

**Discussion:** Each year, a new cohort of up to 70 high school students will be enrolled in MYHealth. We anticipate the MYHealth program will increase interest and persistence in STEM and health research among groups that have been historically excluded in health research careers.

## Introduction

Despite decades of calls for increased diversity in the research workforce, several groups continue to be excluded and thereby underrepresented in Science, Technology, Engineering, and Math (STEM) careers in the United States [1]. In 2019, Black, Indigenous, and People of Color (BIPOC) individuals were awarded only 11.7% of STEM research doctorates, despite making up about a third of the U.S. population and the workforce [1]. Likewise, while women comprised 47% of the workforce, they received only 42.2% of STEM doctorates [1]. Parental education is also associated with postsecondary enrollment, which is an important step on the trajectory toward many STEM professions. College enrollment directly after high school is lower among children whose parents did not graduate high school (50%), attend college (57%), or graduate college (67%), compared to those whose parents have completed a bachelor’s degree (84%) [2]. Low-income youth also continue to graduate high school and enroll in college at lower rates (65% vs. 83%) than their middle- and high-income peers [2]. While these figures have improved slightly in the last decade, significant work is still needed to address these inequities [3, 4].

An initial and persistent interest in science is driven largely by social and environmental factors, making those from historically excluded groups more vulnerable to barriers such as family factors (e.g., attitudes toward science careers, parent interest in scientific activities), pedagogical factors (e.g., teaching culture, classroom culture), extracurricular activities (e.g., science clubs, out-of-school experiences), and social factors (e.g., models of science interest, peer culture), which in turn impact one’s expectations and beliefs that they can succeed in science [5, 6]. High school and adolescence are critical windows to intervene in developing an interest in STEM. This period of the life course is centered on identity development and is when youth begin to understand their skills, interests, and values and begin to make career decisions [6]. When youth are encouraged to practice science and strengthen their beliefs about their efficacy, they are more likely to express interest, achieve, and persist in science. However, this is not the most likely experience for youth from historically excluded groups, who may experience school practices that do not nurture science interests and capacities [2, 7] and have access to fewer out-of-school science experiences, mentorship, and science role models [2, 5].

Despite school requirements for science curricula, some high school students still lack meaningful and relevant opportunities to engage in science and research [8, 9]. Research suggests that teens from historically excluded groups often view school science as “hard” and “discouraging” and do not see science as relevant to their everyday lives [5, 9-11]. Moreover, students may not know any science professionals in their daily lives, making science and research seem abstract. When paired with a lack of advocates, mentors, and support systems, this leaves students from historically excluded groups with more negative experiences of science [11]. On the other hand, when students experience success with science, receive support from parents and other adults, and develop self-efficacy in science, they are more likely to have positive attitudes toward science [8]. Out-of-school experiences, including summer research experiences and after-school programs can help students develop an interest in science, improve science self-efficacy, increase their science knowledge and skills, and build commitment and persistence in high school that may improve enrollment and retention in undergraduate and graduate school programs in the future [12-14]. Critical to the success and effectiveness of programs is exposure to developmentally- and personally-relevant research opportunities, mentorship and social support, and experiences that encourage empowerment, agency, and a sense of purpose [9].

Increasing the participation and success of researchers from historically excluded groups is essential for strengthening innovation and global leadership in health research [15]. Diverse and inclusive scientific teams not only generate novel research questions and methods, but by bringing their life experiences and perspectives, they also offer new problem-solving approaches to persistent issues in research [16]. Furthermore, tackling the innumerable health disparities faced by historically excluded groups requires the support of diverse scientific teams. Thus, the need for a diverse health research workforce serves as a comprehensive theoretical and empirical foundation for the Michigan Youth Health (MYHealth) program [6, 11, 12, 17]. The overall goal of the MYHealth research training program is to address the persistent problem of underrepresentation of historically excluded groups in health research careers. This project combines an innovative, youth-centered research program with a community-based participatory research (CBPR) approach to partner with high school students as co-researchers. The training program aims to help youth build persistence toward health research careers by developing: 1) researcher identities, 2) scientific literacy, 3) scientific self-efficacy, and 4) teamwork and leadership self-efficacy. Engaging high school students in a research training program is advantageous because postsecondary decisions and occupational interests are often crystallized and plans formulated during adolescence [9].

### Conceptual overview

Because students from excluded groups are more likely to encounter external barriers (e.g., fewer STEM role models, less access to rigorous STEM coursework) that are associated with dropping out of the STEM and health research pipeline, it is imperative to understand how to support their persistence toward science careers effectively [18-20]. Based on an ethnically diverse sample, Chemers and colleagues demonstrated that researcher identity, science self-efficacy, and leadership and teamwork self-efficacy mediated the relationship between science support experiences and persistence in science and health research careers [18]. Extrapolating from these findings, researchers have validated the effectiveness of interventions that incorporate a range of out-of-school programs that provide opportunities for research experiences, mentoring, and community involvement [12-14]. Building on Chemers’ framework for understanding persistence in science, MYHealth integrates social cognitive theory, expectancy-value theory of achievement motivation, and CBPR. The MYHealth framework is outlined in Fig 1.

**Fig 1.**
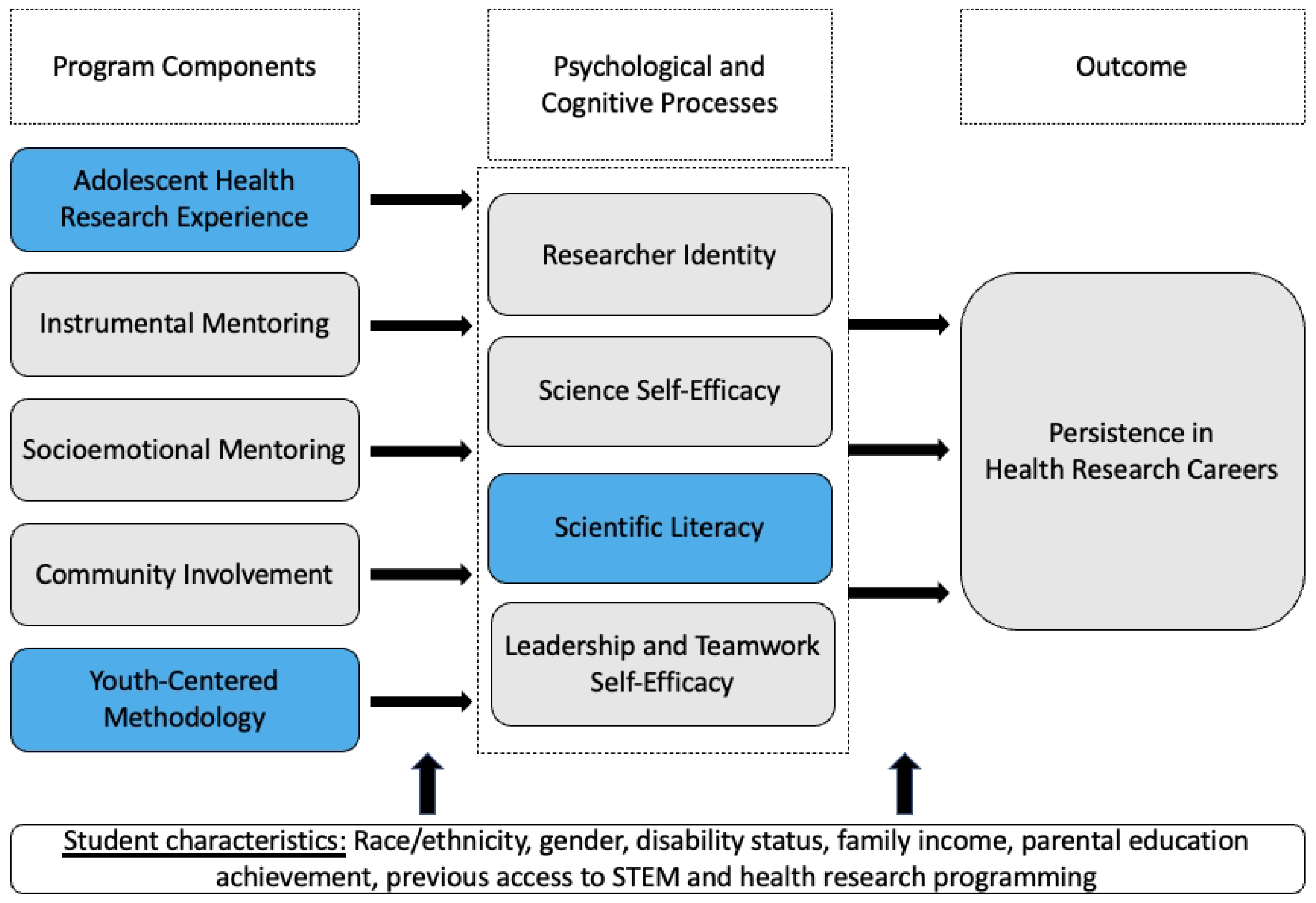
MYHealth conceptual framework, adapted from Chemers et al., 2011.

### Social cognitive theory (SCT)

SCT seeks to explain human learning and motivation and has been applied to domain-specific learning (e.g., science learning) across fields [21-25]. SCT identifies three components that interact to influence learning: personal characteristics (e.g., self-efficacy), environmental characteristics (e.g., school, support, community), and behavior (e.g., pursuit of opportunities). These dynamics have been extensively studied in relation to science interest, achievement, and career choice [18, 26, 27], and among historically excluded youth [28, 29]. Research demonstrates that students from historically excluded groups have fewer opportunities to have their science self-efficacy beliefs nurtured; these dynamics, in turn, suppress motivation and science self-efficacy beliefs and ultimately contribute to fewer youth pursuing science degrees and research careers [30-32].

### Expectancy-value theory of achievement motivation

Linked to SCT, the core of the expectancy-value theory of achievement motivation is that students’ achievement and decisions related to achievement are determined by expectancies for success in that domain and the perceived value of that domain in their lives [33, 34]. According to this theory, achieving and maintaining an interest in science results from students’ beliefs about their likelihood of success in science and the value they assign to science. Applied to MYHealth, expectancy-value theory posits that students from excluded groups can be supported to view science and research careers as attainable and important through opportunities that: 1) increase positive perceptions related to their science and research skills, characteristics, and competencies and 2) increase positive perceptions related to their personal values and goals. Taken together, science- and research-related SCT and expectancy-value theory help us to understand the processes that contribute to persistence in science and research and associated careers, particularly among historically excluded students.

### Community-based participatory research (CBPR)

CBPR is built on core principles that support partnership, capacity building, co-learning, and action toward equity and social justice [26, 35, 36] and has decades of research demonstrating its effectiveness in creating partnerships and enhancing the quality and rigor of research [24, 37-41]. CBPR follows the research cycle and includes building a research team (with nontraditional research partners, like youth), identifying the needs and priorities of a community, asking questions, gathering data, interpreting data from the lens of the community, and implementing action strategies toward health and social justice.

Using CBPR as a model for engaging youth as co-researchers benefits youth directly (e.g., research training, communication skills, problem-solving and critical thinking skills, leadership, preparedness for college), their communities (e.g., community literacy, improved programs and services, improved policy), and research quality (e.g., improved recruitment, retention, sustainability) [42, 43]. Given its overall inclusive and equitable approach, CBPR is a compelling and effective methodology for engaging students from marginalized groups and supporting their long-term interest in science.

In summary, the MYHealth program builds on the theoretical underpinnings of SCT, expectancy-value theory of achievement motivation, and a CBPR approach to create an out-of-school research experience for youth that includes experiences with an applied adolescent health research project, instrumental and socioemotional mentoring from academic researchers and a community of learners, and community involvement through research projects that impact youth and their communities. Through these program components, we will impact four primary psychological and cognitive processes: researcher identity, science self-efficacy, leadership and teamwork self-efficacy, and scientific literacy. By using MYHealth to develop these domains among youth co-researchers, we will achieve our long-term goal of increasing persistence toward health research careers.

### Overview of the MYHealth program

The MYHealth program is a research training program with the long-term goal of increasing the representation of groups traditionally excluded from biomedical, behavioral, and clinical research, including individuals from low-income backgrounds, first-generation college students, and racial and ethnic minority youth. The MYHealth program will enroll 4 successive 12-month cohorts of high school students who self-identify as underrepresented in STEM. The MYHealth program achieves its overall aim via multiple phases each year, as outlined in Fig 2. The Summer Launch, Impact Projects, and Peer Leadership phases each have specific goals, activities, outputs, and outcomes that are summarized in Table 1 and detailed in the sections that follow. The program curriculum was developed through an iterative process of piloting and feedback from high school students from the local population. A condensed curriculum was presented to a pilot cohort of participants and the refined curriculum is detailed below. Each year, modifications will be made based on our formative evaluation.

**Table 1.**
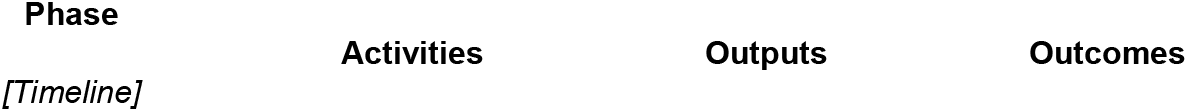
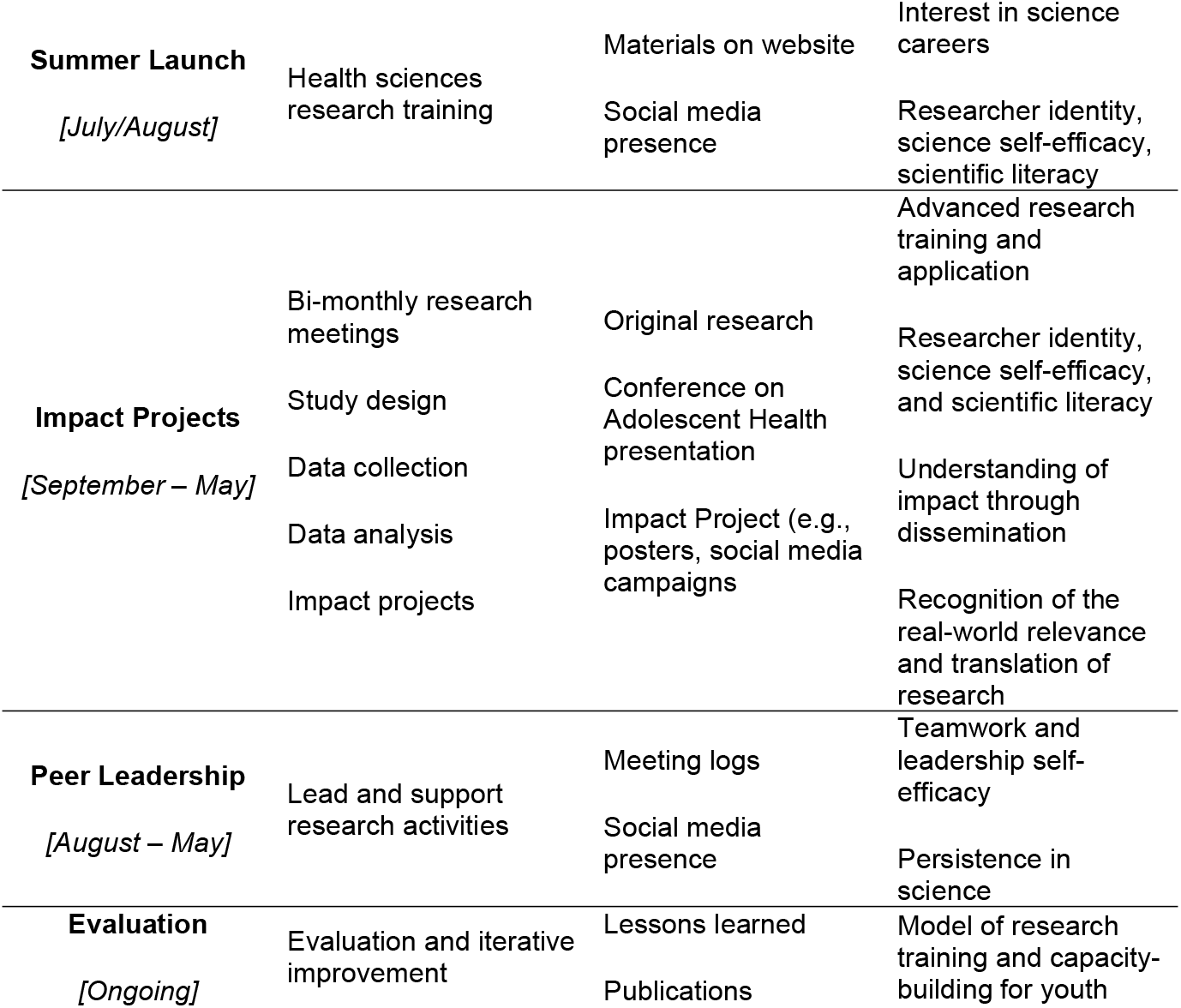
MYHealth program struture.

**Fig 2.**
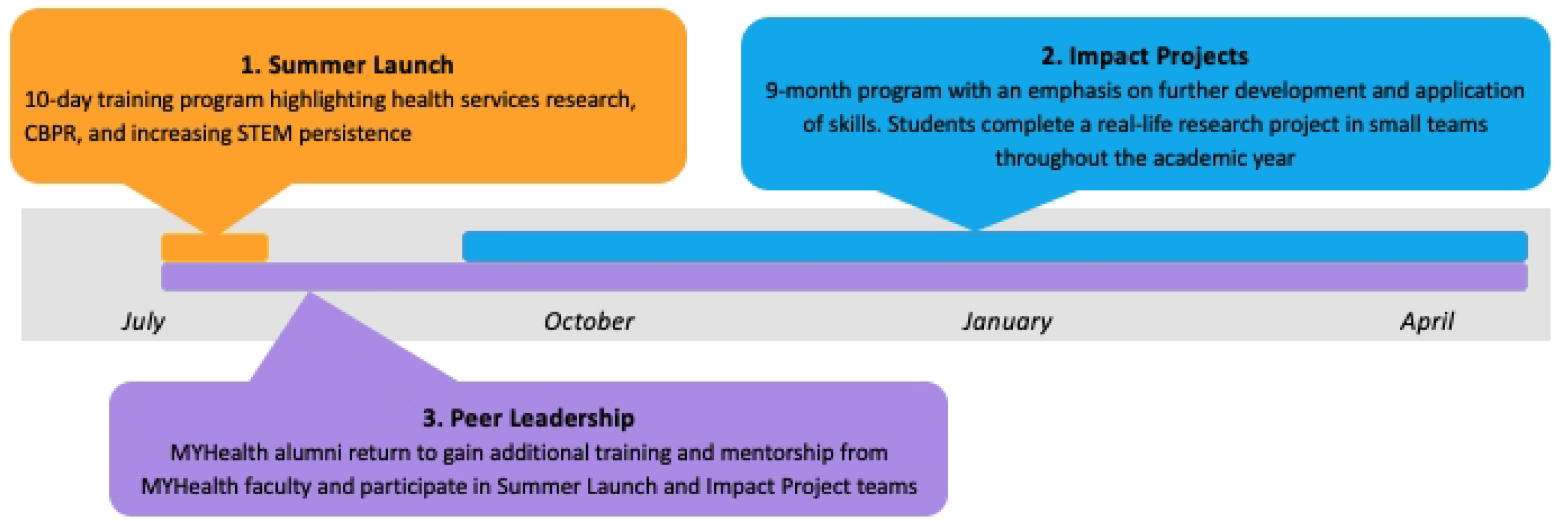
MYHealth program overview.

### Summer launch

The Summer Launch will be a 10-day training in CBPR and health research methodologies for up to 70 students each year. The overall goal is to train high school students to be co-researchers who can work alongside academic researchers on studies of adolescent health to develop their interest in STEM and incubate researcher identities. A sample of Summer Launch curriculum topics and program constructs are outlined in Table 2. The Summer Launch is designed to take place in person on the University of Michigan campus to build excitement and energy related to research, create community among students from different communities in Southeast Michigan, and introduce youth to college campuses. The Summer Launch will be adapted and conducted virtually when meeting in person is not possible due to COVID-19 restrictions.

**Table 2.**
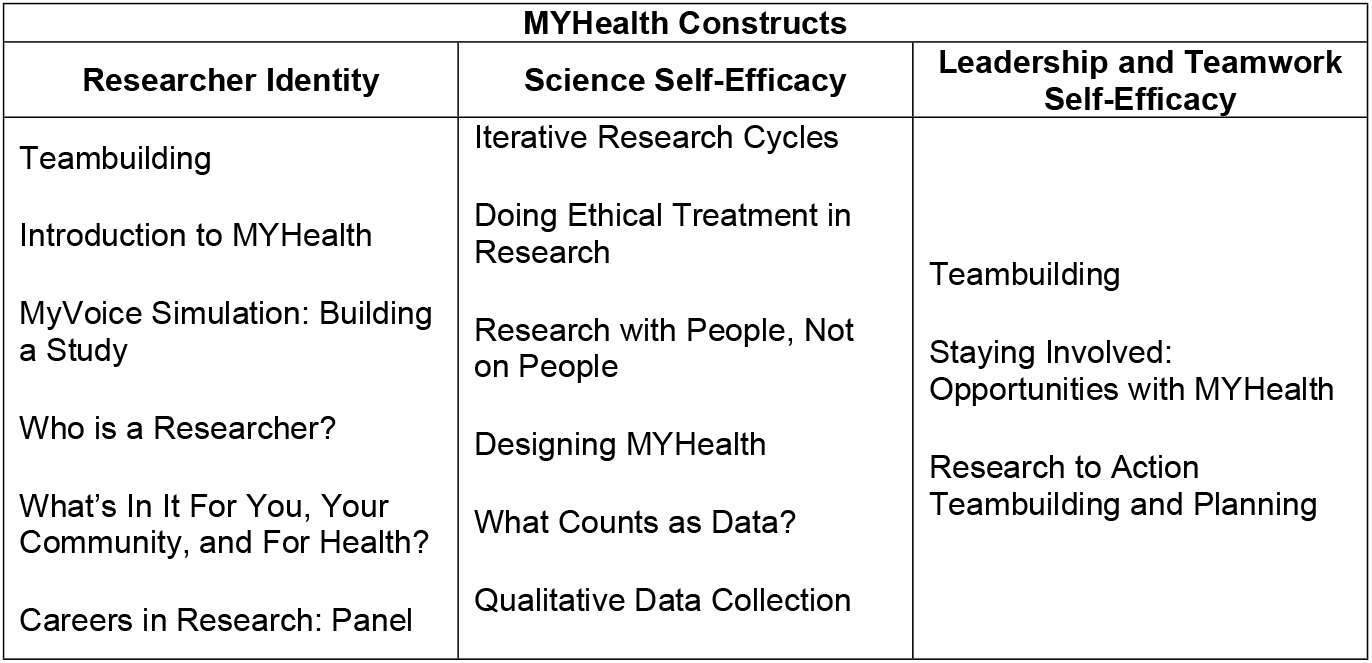
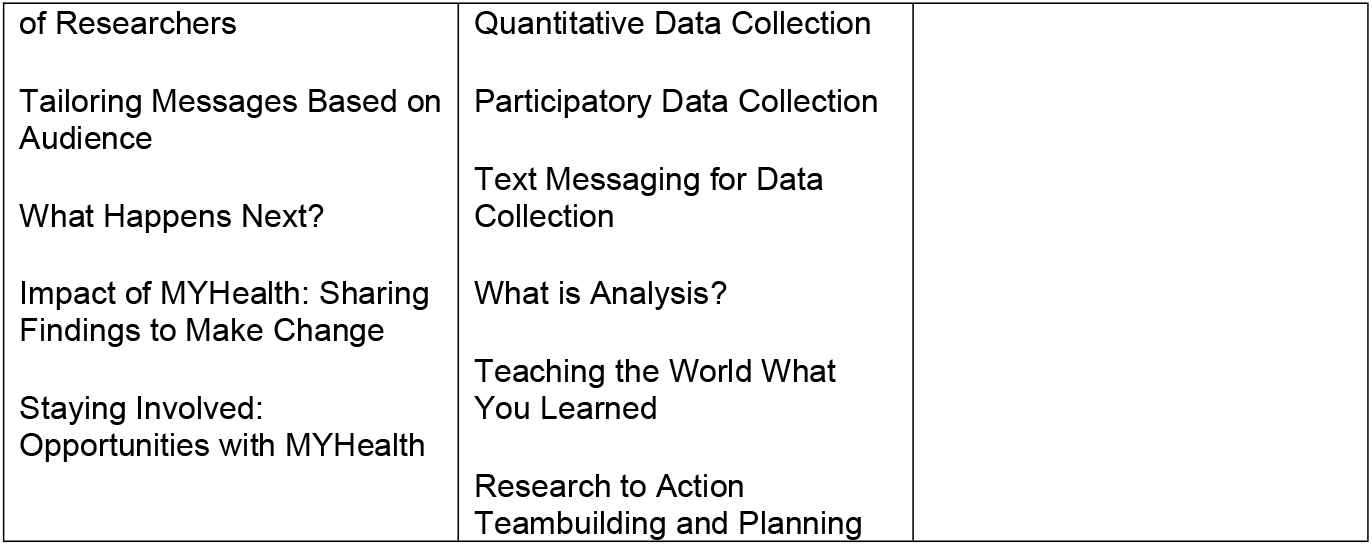
Sample summer launch agenda topics aligned with MYHealth constructs.

Throughout Summer Launch, students will learn about research methods, CBPR, and various adolescent health studies (Table 2). We will create applied learning experiences using MyVoice, a national text message poll of 14-24-year-olds to understand adolescents’ views on a range of health-related topics [44]. For all discussions, MyVoice will serve as a specific data source to illustrate research elements in the context of a real-world project. For example, when learning about data collection, youth will review existing MyVoice question sets and participate in a MyVoice pilot, receiving SMS messages like MyVoice participants. Consistent with our CBPR approach that trains youth to be co-researchers, the Summer Launch agenda and activities will align with equity, inclusion, and social justice principles. Each topic will consider the importance of ethical and responsible conduct of research—from designing a study with the well-being and needs of participants in mind, to obtaining consent, to fairly representing data and disseminating it to all relevant stakeholders.

### Impact projects

At the end of the Summer Launch, all students are invited to apply to participate in the Impact Projects phase, a nine-month program that aims to build on the overall momentum, skills, and interests developed in the previous phase. When applying, students will specify their interest in various adolescent health topics to help guide the direction of the Impact Projects. We anticipate that up to 30 high school students will complete Impact Projects in the following academic year.

Impact Projects are guided by several key principles, including equitable partnership as co-researchers, positioning youth as experts of their own experiences, studying a relevant and meaningful topic to youth, and sharing findings with diverse audiences. By engaging in these collaborative activities, the students will develop their researcher identities, enhance their sense of scientific self-efficacy, and hone their scientific literacy.

Students will meet virtually, approximately twice per month, in small teams that include a faculty mentor, an undergraduate and/or graduate student research assistant, and up to two Peer Leaders who are MYHealth alumni. Projects are conducted virtually to broaden access to MYHealth for high school students across schools and communities. To effectively engage high school students, meetings will be highly interactive, including group processes and applied exercises. See Table 3 for a sample of the Impact Projects agenda. Activities will mimic the research process, including three key phases: 1) Study Design; 2) Data Collection and Analysis; and 3) Dissemination. These phases will include ongoing training in CBPR, health research methods, and the responsible conduct of research. Though most of the training and activities will occur in small groups, each team will also share their progress with other Impact Project teams and program faculty to receive feedback and practice research communication.

**Table 3.** Impact projects agenda topics by stage.

| Impact Project Stages |  |  |
| --- | --- | --- |
| *[Timeline]* |  |  |
| Study Design | Study Design | Study Design |
| *[September – October]* | *[September – October]* | *[September – October]* |
| Identifying a priority area | Identifying a priority area | Identifying a priority area |
| Gathering and summarizing existing literature | Gathering and summarizing existing literature | Gathering and summarizing existing literature |
| Developing a research question | Developing a research question | Developing a research question |
| Writing a research plan | Writing a research plan | Writing a research plan |
| Developing, piloting, and revising open-ended survey questions | Developing, piloting, and revising open-ended survey questions | Developing, piloting, and revising open-ended survey questions |

### Peer leadership

The overall aim of the Peer Leadership phase is to provide ongoing social and academic support, increase motivation, and improve self-efficacy for students from historically excluded groups pursuing STEM and health research careers [28, 41]. Peer Leaders will receive additional coaching and support from program faculty, including additional training in CBPR and health research methodologies. Selection criteria for being a Peer Leader include having an aptitude and interest in becoming a researcher. We anticipate that up to 10 of the 30 participating Impact Project students each year will be Peer Leaders in the following year.

Peer Leaders will interact closely with MYHealth high school students and program faculty during the Summer Launch and Impact Projects. Peer Leaders will be part of Impact Project teams and will provide coaching and guidance to their team members, work alongside faculty mentors, and maintain logs of research activities and progress for the program evaluation. Peer Leaders will be encouraged to contribute to dissemination efforts, including 1) contributing to manuscripts and presentations to share evaluation findings; 2) supporting resource-sharing efforts through in-person presentations to local STEM programs; 3) supporting resource-sharing efforts through the website and social media strategies.

### Methods

A mixed methods approach will be used to collect multiple quantitative and qualitative measures of implementation and outcomes over five years. An explanatory sequential design (quantitative followed by qualitative) will be implemented wherein survey responses will inform final interview questions to evaluate the impact of the MYHealth program on participants’ interest in and persistence in STEM research. The Institutional Review Board at the University of Michigan approved the evaluation study of the MYHealth program (HUM00213914, HUM00214694, HUM00214949). All MYHealth participants will consent or assent to participate in the research and parental consent will be collected for minors. Participants will be assigned a random identifier at the start of the program to protect their anonymity and confidentiality throughout data analysis. Because survey questions are focused on attitudes and beliefs about science and research and are not sensitive in nature, there is minimal risk to the participants throughout the course of the study.

### Setting

The MYHealth program focuses on recruiting high school students based in Southeast Michigan including, but not limited to, the Detroit, Southfield, Flint, East Lansing, Ypsilanti, and the surrounding areas. These communities in Southeastern Michigan will be well-suited for recruitment of high school students from historicall excluded groups and have a clear need for additional science training that is more engaging, relevant, and impactful than traditional classroom science. During the 2016-2017 school year, science proficiency rates in these communities lagged behind other schools in Southeastern Michigan and the state of Michigan. For example, several focal communities had science proficiency rates of 11^th^ grade high school students between 9% and 11%. When looking at economically disadvantaged districts, these numbers dropped below 8%. While state proficiency assessments are not the sole determinant of science interest, enrollment in science courses in college, or future STEM careers [45], high school science achievement can alter youth’s perceptions of their capacity to engage in science [6, 18, 46].

### Recruitment

High school students will be recruited through ongoing partnerships with community partners who have established networks of schools, teachers, and communities and will connect eligible students and families to the MYHealth program. These partners include university centers that serve students and schools, local school districts, and an academic support program. Our community partners will distribute recruitment materials directly (e.g., information sheets, flyers, emails) through their networks. Interested students and their families will be directed to complete an online application form or contact our research team via email or telephone. The application form (administered via the Qualtrics secure platform) asks interested high school students and their families to provide contact information, state their interest in MYHealth, and explain why they think they would be a good fit for the program. Strong applicants will demonstrate an interest in health, community engagement, research, or the capacity to work in teams.

### Eligibility criteria

Incoming high school students living in Southeast Michigan who self-identify as members of an underrepresented group in science or health will be invited to participate. No racial or ethnic groups will be excluded from participating in the study.

### Data collection and outcome measures

Numerous scales that have been previously validated with youth will be used to measure youth experiences with STEM and research [8], researcher identity [38, 40], interest in and persistence in STEM careers [24], scientific literacy [37, 39], and leadership, and teamwork-self-efficacy [18]. Measures were tested during a pilot phase and then revisions were incorporated. Table 4 presents an overview of the key measures and the critical time points at which they are collected.

**Table 4.**
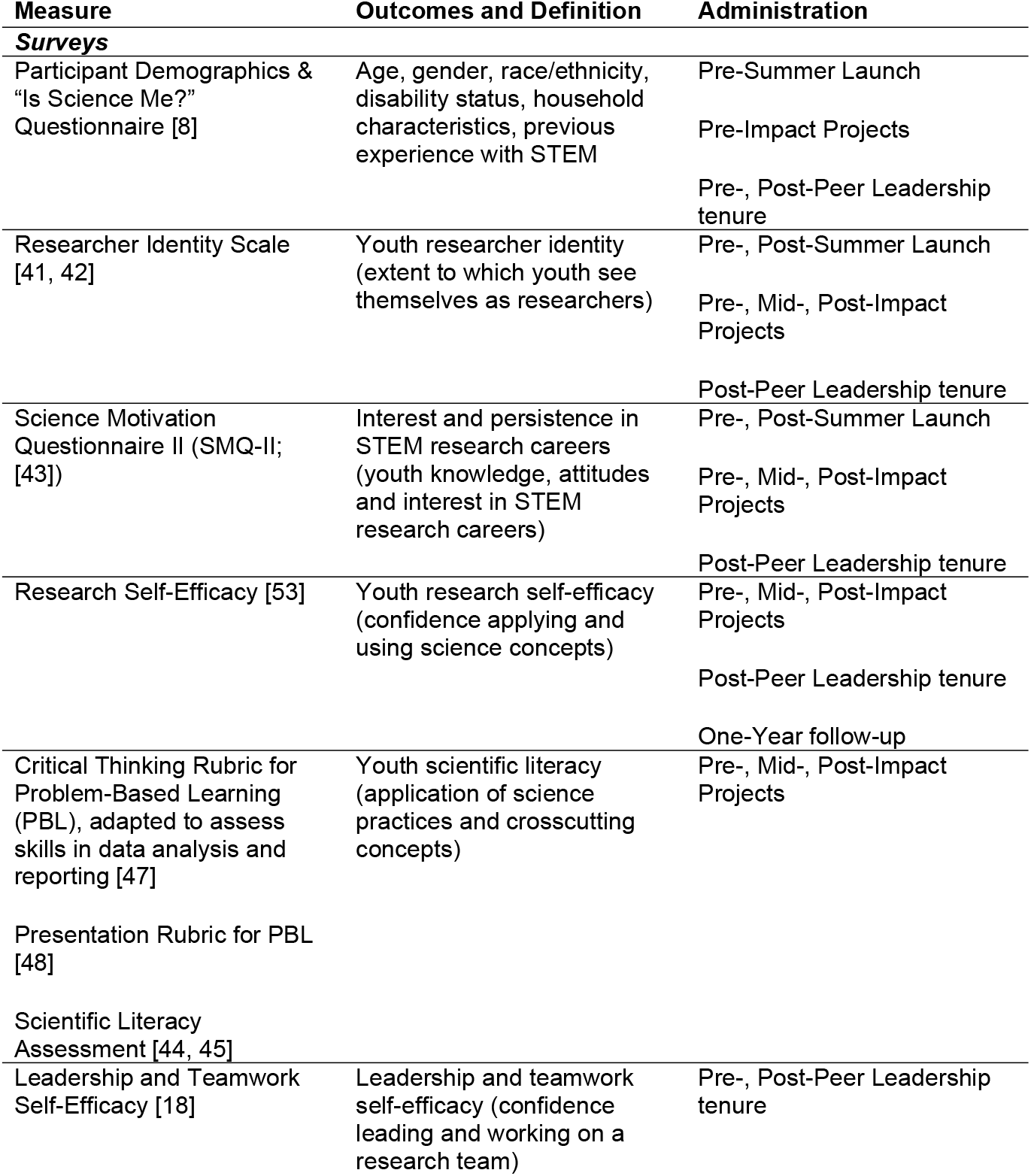
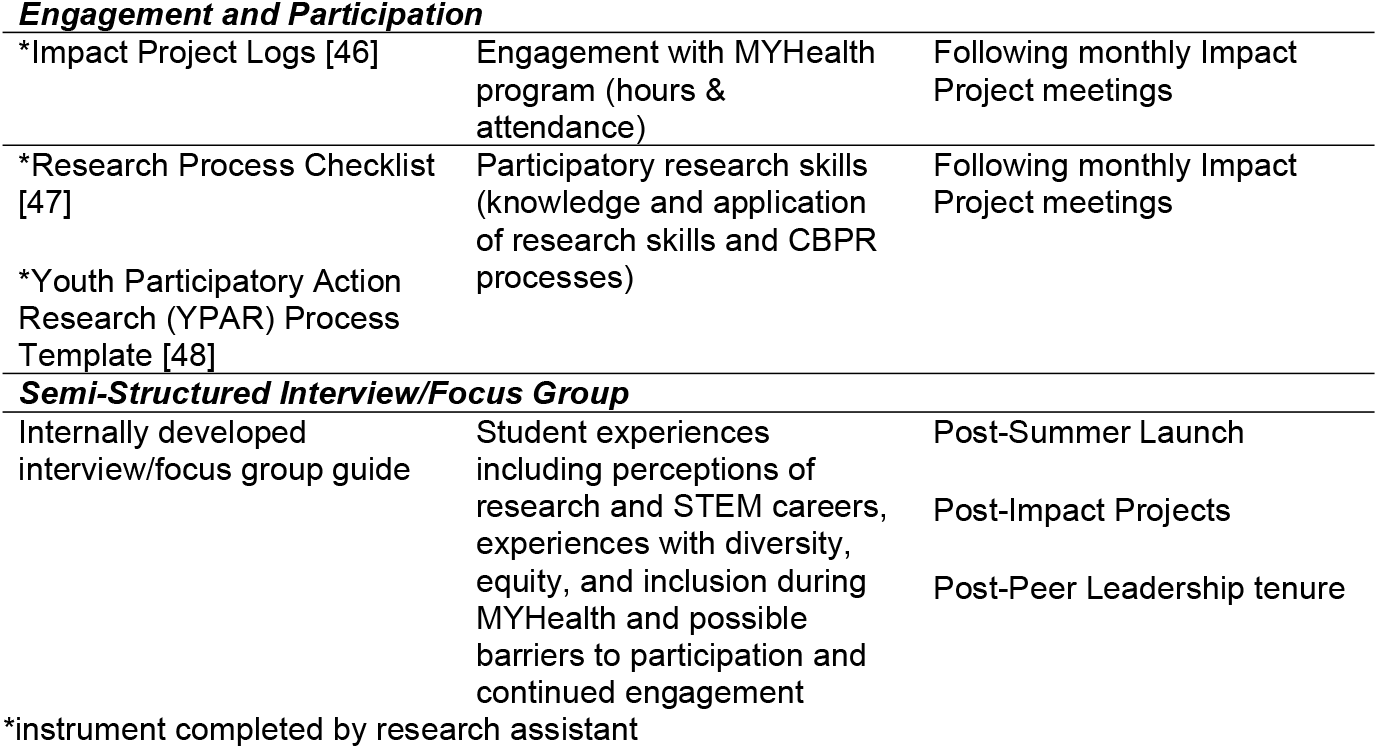
Overview of evaluation instruments and data collection timepoints.

### Surveys

An online survey will be administered online via Qualtrics at multiple timepoints during each program phase, including pre- and post-Summer Launch, pre-, mid-, and post-Impact Projects, pre- and post-Peer Leadership, and one year after completing all program activities (Table 4). The one-year follow-up survey will track youths’ continued interest and persistence in STEM research careers, knowledge and skills related to STEM research, and how the program has influenced their engagement in other research activities.

### Engagement and participation

Each year students’ reactions to the Summer Launch and the Impact Project programs will be collected to make ongoing course improvements. In addition, we will design Peer Leader and graduate research assistant logs to facilitate reflections about what worked in each meeting and what could be improved before the next session for both the Summer Launch and the Impact Project programs.

A research assistant will track program hours and attendance to gauge dosage [47] in the Impact Projects. Logs will be used to track the implementation of CBPR processes at monthly meetings. The logs will consist of a *Research Process Checklist* [48] to understand progress in the research design process (e.g., stated research problem, defined variables, and specified procedures to analyze data). Finally, the graduate research assistant will complete an observational rating scale, *Youth Participatory Action Research Process Template* [49], to track CBPR processes demonstrated by the Peer Leader and Impact Project students in each small group, including training and practice of research skills, promoting strategic thinking, group work, networking, communication skills, and power sharing. In addition, Evaluators will work with the graduate research assistants to complete the *Critical Thinking Rubric for PBL [50]* at three points during the academic year. In the spring of each year, Impact Project students will disseminate research findings. The graduate research assistant will use the *Presentation Rubric for PBL* to capture how students are communicating research processes and findings with youth, community, policy, and academic audiences [51].

### Semi-structured interviews and/or focus groups

Each year at the end of the Summer Launch, up to 10 students will be invited to participate in a semi-structured interview or focus group [52]. We will use a semi-structured interview guide that explores student experiences during the Summer Launch, including perceptions of research and STEM careers, experiences with diversity, equity, and inclusion during MYHealth, and possible barriers to participation and continued engagement. Interviews will be recorded and transcribed.

Similarly, at the end of the Impact Project each year, a sample of up to 10 students will be asked to participate in semi-structured interviews or a focus group to understand processes that occurred during implementation with each cohort. Questions will probe topics such as students’ perceptions of STEM and health research careers, their experiences with diversity, equity, and inclusion on the Impact Projects, and possible barriers to participation and continued engagement. All interviews will be recorded and transcribed for analysis.

### Analyses

Overall, there are parallel strategies to evaluate all three program components: the Summer Launch, Impact Projects, and Peer Leadership. A Repeated Measures Analysis of Variance will be conducted to analyze the quantitative items in the pre-, post-, and follow-up surveys. Small-scale analyses will be performed on each program phase, and a longer-term evaluation will be conducted on longitudinal data. Over time, between-subjects factors may be added to account for differences between cohorts, or disaggregate data by demographic factors (e.g., gender, ethnicity). The final reporting will utilize appropriate statistical significance and associated effect sizes.

Interview data will be analyzed with an inductive Outcome Harvesting approach [54, 55], in which stakeholders identify individual and group impacts that mattered most, and speculate on program elements that led to these outcomes. Codes will be applied to all interview transcripts and then synthesized into related themes and conclusions. Findings will be used to inform future years of the MYHealth program.

## Results

We anticipate that up to 280 total high school students from historically excluded groups will be enrolled in MYHealth across four years: up to 280 will have completed the Summer Launch, a subset of up to 120 will have completed the Impact Projects, and a smaller subset of up to 40 will have become Peer Leaders. Primary quantitative outcomes for this evaluation are Researcher Identity and STEM research career interest. Our power analyses indicate that we need 90 students each in the Summer Launch and Impact Projects to detect a minimum effect size of 0.3 which is considered small by conventional interpretations [56]. This assumes 80% power at 5% Type 1 error (two-tailed t-test). Given our anticipated sample, we will have sufficient students to make conclusions about changes in Researcher Identity and STEM career interest even with attrition.

For the qualitative components of the program evaluation, we will ensure that we have a sufficient sample size to achieve thematic saturation. In qualitative approaches, thematic saturation refers to the point at which there is adequate evidence to develop themes and no new insights are gathered with continued data collection. For interview studies, thematic saturation is often estimated to be achieved between 6-20 interviews [57, 58].

## Discussion

MYHealth is limited by our small cohort size and geography. The cohort size for each phase is limited to ensure that students receive adequate time with faculty mentors and support for their small group projects. Over the four years of the project, we will be able to understand the impact of the research training program. Currently, only students from Southeast Michigan can participate. If successful, future iterations of MYHealth could leverage our virtual program to extend to another setting outside of Southeast Michigan.

The first complete year of the MYHealth program began in the Summer of 2022, and will re-open annually for the following four years. We anticipate data will become available through peer-reviewed publications beginning in the first two years and on several more instances throughout the course of the project.

## Conclusion

The MYHealth program capitalizes on Social Cognitive Theory, Expectancy Value Theory, and a CBPR approach to design and implement a program that aims to address the continuing lack of diversity within the STEM workforce. The program assesses the role and intersection of personal characteristics (e.g., researcher identity, self-efficacy), social and environmental factors, and behavioral influences to spark and maintain interest in STEM careers.

## Data Availability

No datasets were generated or analysed during the current study. All relevant data from this study will be made available upon study completion.

## Authors’ contributions

All authors, MD, JR, AA, TC, MDi, KH, LMV, DCW contributed equally to the the work by conceptualizing the study aims and writing, reviewing and editing the study protocol.

